# Gaze-control training in a sample of inattentive young adults: A Confidence-in-Concept study of neural mechanisms

**DOI:** 10.1101/2024.11.17.24316746

**Authors:** Alice E. Waitt, Jyothika Kumar, Lauren Gascoyne, Bryony Waters-Harvey, Abdulrhman Shalabi, Jacob Habgood, Peter Collins, Maddie Groom, Peter F Liddle, Elizabeth Liddle

## Abstract

**Background:** Mounting evidence links attentional disorders to impairments in oculomotor control. Moreover, the brain’s oculomotor control network forms the backbone of neurocognitive systems typically impaired in neurodevelopmental conditions like Attention Deficit/Hyperactivity Disorder (ADHD). RECOGNeyes is an eye-tracker controlled computerised cognitive training game designed to improve oculomotor control. In this confidence-in-concept study, we investigated the effects of RECOGNeyes training on oculomotor control and its neurological correlates in an inattentive sample of young adults.

**Methods:** Thirty-five participants receiving support for ADHD, dyslexia or a related condition, were randomised to two, three, or four RECOGNeyes training sessions per week, at home, for two weeks. Total training time was recorded. Outcomes included measures of reading efficiency, and performance on an antisaccade task, during which brain oscillations were recorded magnetoencephalography. Electrophysiological measures included anticipatory alpha-band oscillatory amplitude in the frontal eyefields, and anticipatory beta-band amplitude in dorsolateral pre-frontal cortex. Oculomotor network connectivity at rest was measured using functional magnetic resonance imaging.

**Results:** Antisaccade task performance and oculomotor indices of reading efficiency improved after training, regardless of achieved training time. Greater total RECOGNeyes training time was associated with greater improvements in antisaccade performance; reduced fixation durations during reading; and greater anticipatory reductions in FEF alpha and DLPFC beta that were also associated with reduced antisaccade reaction time cost. Greater training time was also associated with reduced between-hemisphere resting-state connectivity, and increased within-hemisphere connectivity in the left hemisphere.

**Conclusions:** We found evidence that RECOGNeyes gaze-control training improves oculomotor control, with possible transfer to reading efficiency, as well as associated changes in electrophysiological correlates of anticipatory attention. Resting state connectivity changes suggest plastic changes within the oculomotor network in the direction of increased hemispheric independence. Next steps will be to establish whether the effects of RECOGNeyes transfer to real-world benefits in children with neurodevelopmental conditions like ADHD.

**Funding statement:** This project was funded MRC Confidence in Concept award (Grant ID CiC2017026); Wellcome Seed Award (202122/Z/16/Z); an MRC PhD studentship (AW); and a PhD studentship funded by Institute of Mental Health (PC).

**Competing Interest Statements:** PC and EL have a revenue-sharing agreement with the University of Nottingham regarding inventors’ share of any revenue accruing to the University arising from future commercialisation of the RECOGNeyes.

**Author Declarations:** I confirm all relevant ethical guidelines have been followed and all necessary ethics committee approvals have been obtained.

## Introduction

Cognitive control is “the ability to regulate thoughts and actions in accordance with internally represented behavioural goals” (1). Cognitive control brain networks typically develop as children mature (2). However, maturation of cognitive control may be delayed in some neurodevelopmental conditions, including Attention-Deficit/Hyperactivity Disorder (ADHD). ADHD is characterised by difficulties with control over attention and action that can persist into adulthood, even when symptoms are substantially remitted (3).

Control over gaze direction is closely linked to *attentional* control (4); we tend to pay attention to what we are looking at. Furthermore, resisting visual distraction involves inhibitory *motor* control over reflexive eye saccadic eye movements (5). Gaze-control therefore sits at the interface between control over attention and action, and gaze-control tasks are commonly used to evaluate cognitive control (6).

One such task is the antisaccade task, which involves inhibiting an automatic saccadic eye-movement towards a peripheral stimulus, and instead making an “antisaccade” in the opposite direction (6). Two meta-analyses of studies of oculomotor performance in ADHD provide substantial evidence for impaired antisaccade performance (7,8). Maron et al. (8) found particularly large effect sizes for impairments in saccadic inhibition and fixation control.

The brain’s oculomotor network includes: visual cortex; frontal eyefields (FEF); parietal eyefields (PEF); cortical regions involved in attention, such as the dorsolateral prefrontal cortex (DLPFC) and anterior insula; subcortical and midbrain structures; the cerebellum. In their review of oculomotor performance in ADHD, Maron et al. (8) cite evidence that the brain’s oculomotor network forms the backbone of the networks that underpin the cognitive impairments typically impaired in ADHD, including selective attention and set-shifting, inhibitory control, and working memory.

Several other neurodevelopmental conditions have substantial rates of co-occurrence with ADHD (9–15), including Specific Learning Disabilities (SLDs), Reading Disorder/dyslexia, Developmental Coordination Disorder (DCD) and autistic spectrum disorder (ASD). Oculomotor control impairments have also been found in these conditions, suggesting that oculomotor control dysfunction may be a common factor (9,12,12,14,15,15–19).

In a typical antisaccade task, a pre-stimulus cue allows the participant to prepare to make either a prosaccade or an antisaccade in response to the upcoming peripheral target. Electrophysiological evidence indicates that for antisaccade trials, this anticipatory period is associated with coupled increases in alpha-band power in the FEF, and beta-band power in the DLPFC (20). Alpha-band oscillations are associated with attentional inhibition (21), and beta synchronisation with top-down control (22). These electrophysiological correlates of antisaccade responses appear to develop over adolescence (23), suggesting that they index maturing cognitive control capacity, and, specifically, the capacity to control direction of gaze.

Beta oscillations in motor cortex typically desynchronise in preparation for a movement, then rebound to above-baseline levels following movement completion, an effect known as Post-Motor Beta Rebound (PMBR) (24). PMBR is more marked in adults than children, and is reduced in some developmental conditions, including ADHD (25,26). PMBR appears to index confidence in the motor prediction and a return to the “status quo” (27,28), and has been observed in complex saccadic tasks as well as limb movements (29).

Attentional control may also be impacted by arousal dysregulation (30). Pupillometry can be used to measure both phasic arousal in response to stimuli, as well as tonic levels of arousal (31). We postulated that pupillometric arousal measures would be associated with measures of oculomotor performance, and potentially impacted by gaze-control training.

We also postulated that suboptimal oculomotor control might be reflected in immature intrahemispheric and interhemispheric patterns of functional connectivity between nodes of the oculomotor network, as measured by resting state Magnetic Resonance Imaging (rsMRI). Functional connectivity tends to be predominantly interhemispheric at birth, and shift towards an intrahemispheric predominance over development, with the development of greater hemispheric independence and specialisation (32,33). Consistent with the hypothesis of delayed maturation in ADHD, interhemispheric homotopic connectivity has been shown to be greater in ADHD than in typically developing participants (34–36)

### Oculomotor control training

RECOGNeyes is a computer game developed by authors Collins, Liddle & Habgood to improve oculomotor control in ADHD and related conditions. A laptop-mounted eye-tracker allows players to control gameplay through changes in gaze-direction. RECOGNeyes includes six mini-games, each designed to train different aspects of oculomotor control capacity, including selective attention, motor inhibition, motor timing, and spatial working memory:

- Maintaining fixation in the face of visual and auditory distractors.
- Cancelling a planned saccade.
- Making an antisaccade.
- Executing a saccade within a learned time window.
- Monitoring the peripheral vision for a saccade target.
- Delaying a saccade to a location until a stimulus appears at a different location.

Task difficulty adjusts to the player’s skill level: adaptive trial-by-trial adjustments to time-limits maintain rates of success at motivating levels, while skill consolidation opens access to more challenging activities.

### Study Aims and Objectives

We conducted a confidence-in-concept study to evaluate the impact of RECOGNeyes training on oculomotor performance and brain networks involved in cognitive control in a sample of inattentive young adults. We randomly assigned training schedules of different intensities in order to link changes in oculomotor performance and neural oculomotor control measures with the intensity of training undertaken.

We predicted several measures on which RECOGNeyes could have an impact, and on which the degree of change would be correlated with training intensity:

1. Oculomotor Performance

- Performance on a trained (antisaccade) task as measured by saccade latencies and accuracy
- Near transfer to an untrained oculomotor task: Eye movement efficiency during reading
- Far transfer to reading scores on the TOWRE-2 reading efficiency test.
2. Electrophysiological oscillatory power for antisaccade trials:

- Anticipatory period:

- Beta-band (13-30 Hz) power in DLPFC
- Alpha-band (8-12 Hz) oscillatory-power in FEFs
- Coupling between beta-power in right DLPFC and alpha-power in FEFs
- Post target:

- Increased ERD and PMBR in FEFs
3. Pupillometric measures of phasic and tonic alerting:

- Phasic changes in response to task stimuli
- Tonic pupil size
4. Oculomotor resting state connectivity between oculomotor network nodes, assessed using rsMRI:

- Intrahemispheric connectivity
- Interhemispheric connectivity between homotopic nodes

## Materials and Methods

The study protocol was reviewed and approved by the Faculty of Medicine and Health Sciences (FHMS) Research Ethics Committee, University of Nottingham (Ethics Reference Number: 154-171).

### Experiment design and data acquisition

#### Participants

Thirty-five young adults (20 female) aged 19-31 (*M* = 24) seeking support for suspected or confirmed ADHD or an SLD (dyslexia, dyscalculia, DCD) were recruited, primarily through University of Nottingham Academic Support Services. Exclusion criteria were: MEG or MRI contraindications; partial sightedness or visual field deficit; involvement in other neuroimaging studies during the last 3 months.

#### RECOGNeyes program and training randomisation

Participants were randomised (stratified by age and gender) to undertake two (N=11), three (N=12), or four (N=12) sessions of RECOGNeyes training at home, for two weeks. Sessions were to last from 20-to-30 minutes, and to be spaced out evenly over the week. These instructions were designed to yield a continuous measure of Total Training Time (TTT) across participant. Investigators undertaking the pre-/post-intervention assessments remained blind to participant assignment until the end of the study. TTT was computed from game logs.

Participants attended the research lab twice, before and after RECOGNeyes training. On Day 1, after providing written informed consent, and completing a Magnetic Resonance (MR) safety form, participants disclosed details of any ADHD or SLD diagnoses and current treatments or therapies, and completed the self-report Conners’ Adult ADHD Rating Scale (CAARS) (Short Version) (37). On both Day 1 and Day 2, they performed an antisaccade task in a MEG scanner, underwent anatomical and resting-state functional MR scans, and were assessed on all other outcome measures.

### Outcome measures

Current mental health was assessed using the 12-item version of the General Health Questionnaire (GHQ12) (38).

Single-word reading was assessed using the Test of Word-Reading Efficiency (TOWRE-2) (39), which assesses single-word reading speed. It yields three age-normed standard scores: sight-word efficiency (SWE), based on reading real words; Phonological Decoding Efficiency (PDE), based on reading pseudo-words; and Total Word Efficiency (TWE), based on both. Alternate word-list forms were randomly counter-balanced across participants.

To assess eye-movement efficiency during reading, eye movements from both eyes were recorded using an EyeLink® 1000 Plus eye-tracker (SR Research Ltd.) at 500Hz, while participants silently read three short passages over a period of ∼5 minutes. This was done in the MEG scanner, prior to the antisaccade task. After each passage they answered questions on the content to ensure comprehension. The passages were based on reading material designed for a reading age of 10, matched for complexity. Sets of passages were counterbalanced across participants.

Eye movement reading metrics, derived from Rayner et al. (40,41) were:

- Preferred landing position (PLP): position in word where the first fixation is made (typically 2-3 letters from start
- The mean length (number of letters) of skipped words. In typical reading only short words are skipped.
- Mean proportion of right-to-left (regressive) saccades as a proportion of total saccades.
- Mean duration of fixations (typically shorter in more fluent readers).

### Antisaccade task

The task consisted of 20 pairs of task blocks (6 trials per block), comprising a “prosaccade” block, in which participants had to make a saccadic eye-movement towards a peripheral stimulus, or an “antisaccade” block, in which they had to make a saccade in the opposite direction. Task-order was counterbalanced across pairs. A 15 second rest period between the two blocks of each pair, and a 30 second rest period between each pair was included to facilitate eye-tracker drift correction, and to enable stable MEG covariance estimates.

Participants practiced the task until they could correctly perform 8 prosaccade and 8 antisaccade trials. The task was programmed in EyeLink® Experiment Builder (SR Research Ltd.) and presented via a mirror-projection display.

### MEG and autonomic data acquisition

MEG data was acquired with a 275-channel MEG CTF system (CTF, MISL, Coquitlam, BC, Canada) contained in a three layer magnetically shielded room (MSR), operated in third-order synthetic gradiometer configuration. Sampling rate was 600 Hz with a low-pass anti-aliasing filter at 150 Hz. Participants sat upright during acquisition and were asked to keep as still as possible. Head motion was continuously monitored via energising three head position indicator coils attached at the nasion and preauricular points. A 3D digitiser (Polhemus Inc, Colchester, VT) was used to generate a head shape representation relative to fiducial markers for co-registration of anatomical MR scan to MEG sensor geometry.

### MRI data acquisition

MR images were acquired using a Philips Achieva 3 Tesla (3T) MR system (Philips Medical Systems, Best, The Netherlands).

T1-weighted anatomical images for MEG source localisation were acquired using a standard MPRAGE sequence protocol: 1 mm isotropic resolution, 256 x 256 x 160 matrix, echo time/repetition time (TE/TR) of 2.2/4.5 ms, short interval of 3000 ms, 8° flip angle and SENSE factor 1 for image registration.

rsMRI: Participants observed a fixation-cross for five minutes, eyes open, while echo-planar images (EPIs) were acquired using a 32-channel head coil with SENSE factor 1 in the anterior-posterior direction. Time-to-Repeat (TR) was 2000 ms; 150 volumes were acquired as 32 contiguous axial slices in descending order, with a slice thickness of 3.5 mm, in-plane resolution of 3 x 3, flip angle of 85°, and a field of view of 240 x 240 x 112 mm.

### Data processing and analysis

#### Eye-tracking metrics during reading

Eye-tracking metrics in the pro/antisaccade task and reading task were extracted and computed using EyeLink® Data Viewer (SR Research Ltd.) and processed using custom interactive MATLAB R2021a (MathWorks Inc.) scripts.

#### Antisaccade performance

Saccadic Reaction Time (SRT) was defined as time between target and saccade onset. SRTs of less than 100ms were discarded in order to exclude reflexive express saccades (42). Responses with the first saccade in the correct direction were deemed correct if the SRT was greater than 100ms, and less than 1000ms.

Typically antisaccade SRTs are slower than prosaccade SRTs, indicating an efficiency “cost” for antisaccades incurred by the greater need for oculomotor inhibition (43) of the automatic prosaccade. Antisaccade Cost was quantified for each participant for each day by subtracting their median prosaccade SRT from their median antisaccade SRT.

Accuracy was quantified by computing a discriminability measure, d′ (44), which penalises high antisaccade accuracy if achieved at the cost elevated error rates on prosaccades. Correct antisaccades on antisaccade trials were treated as “hits”; antisaccades on prosaccade trials as “false alarms”.

#### Pupillometry and vergence

Blinks and other artefacts were identified visually and removed using a custom interactive MATLAB R2021a (MathWorks Inc.) script. Missing segments replaced using a spline interpolation (for further details see Supplementary Materials). We computed median pupil dilation rates for each participant during the cue-target period and from target onset to 100ms later. Tonic pupil size was estimated as the median pupil size at cue onset across each trial type. We were unable to compute vergence measures due to frequent occlusion of the eyetracker’s view of one eye by the MEG dewar.

#### MEG data analysis methods

MEG data pre-processing was conducted using DataEditor software (Release: 5.2.1-linux-20060623, VSM MedTech Systems Inc., Coquitlam, BC, Canada). DC offset, 1-150Hz band-pass filter and synthetic 3^rd^ order gradiometer noise cancellation were applied. Data were segmented into 3 second epochs beginning 300ms prior to each cue. Incorrect trials and trials in which head motion exceeded 2.5mm from the starting position were removed. Trained team members visually inspected trials to manually remove any remaining trials containing artefacts from movement or blinking. Source localisation with structural MRIs were performed using FieldTrip toolbox functions (45) with customised MATLAB scripts based on O’Neill et al. (46). A scalar linearly constrained minimum variance beamformer (47,48) was used to derive a spatial filter from the segmented data for each of nine Virtual Electrodes (VEs) (Figure 7A), using a single-shell forward model (49), and a heavy regularisation parameter of 1% of the difference between maximum and minimum covariance to ensure that VE timecourses would be representative of the area surrounding each ROI. These spatial filters were then used to generate VE timecourses for each trial type on each day (50), resulting in nine sets of four virtual electrode (VE) timecourses for each participant.

As our primary hypotheses related to brain oscillations in FEF and DLPFC, for each participant, VE timecourses for left and right FEF and DPLFC ROIs for each day were re-segmented into three-second trial epochs and concatenated across days. For each concatenated VE, Time-Frequency Spectrograms (TFS) were computed using the Continuous Wavelet Transform function from the MATLAB Signal Processing Toolbox, and expressed as amplitudes by taking the absolute value of the complex valued TFS. Forty-four frequency bands between 2 and 40 Hz were each mean-centred, using mean amplitudes evaluated across both days and trial types, then reshaped into 3D matrices representing every 3 second trial, sorted by trial-type and day.

For the anticipatory cue-target interval, ROIs from each hemisphere were analysed separately. To investigate coupling between DLPFC and FEF ROIs during this period, for each hemisphere, a correlation matrix was computed between the TFS amplitude values for FEF and DLPFC ROIs. For post-target data, data from each hemisphere was recategorized as either “ipsilateral” or “contralateral” to the direction of the executed outward saccade, and both target-locked and response-locked TFS matrices computed.

### Resting state functional MR

Ten ROIs (five in each hemisphere: DLPFC, FEF, Anterior Insula, PEF, V1) were defined using the coordinates illustrated in Figure 7. Intrahemispheric connectivity was computed between ROIs within each hemispheres and interhemispheric connectivity between left-right homotopic ROI pairs.

Connectivity change values were computed by subtracting Day 2 values from Day 1 values. To evaluate effects of TTT on connectivity changes while controlling for baseline effects, first, the connectivity change values for each ROI pair were regressed on baseline connectivity for that pair. For effects of TTT on change in connectivity, the residuals from these regressions were analysed using Mixed Linear Models, with TTT as a between-subjects predictor. For homotopic ROI pairs, ROI pair (5 levels) was the single within-subjects factor, while for intrahemispheric ROI pairs, there were two within-subjects factors: intrahemispheric ROI pair (10 levels) and hemisphere (2 levels). A Mixed Linear Model was also conducted on Day 1 intrahemispheric connectivity values to evaluate between-hemisphere intrahemispheric connectivity differences at baseline.

### Statistical analyses

As an exploratory, confidence-in-concept investigation, this study was not powered to find effects of pre-specified size, or to control for multiple predictions. Therefore, following the second recommendation by Benjamin and Berger (51), we provide both *p* value and upper Bayes Factor Bound (BFB) on the odds in favour of the alternative to the null hypothesis, for each statistical test. Upper BFBs were calculated from *p* values using the sample-size adjusted method proposed by Held & Ott (52), and interpreted using their proposed strength-of-evidence descriptors.

#### Baseline characterisation

One-sample t-tests were used to test compare mean CAARS T scores with the population mean of 50, and mean baseline TOWRE-2 scores (SWE, PDE, TWE) to the population mean of 100.

#### Impacts of RECOGNeyes training

Paired t-tests and/or repeated measures ANOVAs were used to evaluate changes in outcome measures between Day 1 and Day 2, regardless of TTT. Changes between Day 1 and Day 2 were then correlated with TTT in order to evaluate the impact of TTT on changes in outcome measures, controlling for baseline performance.

For analyses of MEG time frequency matrices, t tests and correlations were conducted at each time-frequency cell of each matrix, across participants. One-sample t-tests were used to test for deviations from the participant’s baseline, at each cell for each condition and day, and paired-sample t-tests used to test for differences between trial-types and changes between days. Matrices representing between-day differences (Day 2 – Day 1 TFS matrices) were computed and each cell correlated with TTT. In addition, correlations between TFS matrix cells and changes in Antisaccade Cost were computed to evaluate the neural correlates of changes in performance. A nonparametric cluster-based permutation test (53) was used to identify contiguous clusters of TFS cells with *r* or *t* values meeting p<.05 and compute the probability of such clusters under an empirical null distribution.

Similarly, for resting state functional connectivity values, Day 2-Day 1 connectivity change values were correlated with TTT, controlling for baseline connectivity by first regressing change values on baseline values, and using the residual values as baseline-adjusted measures of connectivity change.

## Results

### Participant sample

The final participant sample size was 35 (20 female), aged between 19.6 and 31.1 years (*M*=24.0, *SD*=3.1) years. The most reported diagnosis was dyslexia (N=28: 80%). Six participants reported a confirmed history ADHD. Mean CAARS scores indicated Inattention/Memory symptoms above the population average, but below average levels of Impulsivity/Emotional Lability. Mean TOWRE-2 scores indicated below average SWE and TWE, but not below average PDE (Table 1).

**Table 1:** Summary statistics for CAARS and TOWRE scores. Abbreviations: BDV= Bayes Factor Bound; SWE= Sight-Word Efficiency; PDE= Phonemic Decoding Efficiency; TWE= Total Wordreading Efficiency.

|  | Mean (SD) | Deviation from pop. mean in SDs | t(df) | p | Upper BFB for H1 | Max Strength of evidence for H1 |
| --- | --- | --- | --- | --- | --- | --- |
| *CAARS T scores (population mean=50, SD=10)* |  |  |  |  |  |  |
| Inattention/Memory | 59.88 (10.40) | 1.0 | 5.37 (31) | <.001 | >300 | Decisive |
| Hyperactivity/Restlessness | 51.68 (12.12) | 0.2 | 0.81 (33) | .426 | 1 | - |
| Impulsivity/Emotional lability | 45.52 (8.26) | -0.4 | -3.12 (32) | .004 | 17.3 | Substantial |
| Problems with Self-Concept | 50.57 (8.52) | 0.1 | 0.40 (34) | .694 | 1 | - |
| ADHD index | 52.63 (9.15) | 0.3 | 1.70 (34) | .098 | 1.6 | Weak |
| *TOWRE-2 standard scores at baseline (population mean = 100, SD=15)* |  |  |  |  |  |  |
| SWE | 89.23 (12.85) | -0.7 | -4.96 (34) | <.001 | >300 | Decisive |
| PDE | 98.43 (15.78) | -0.1 | -0.59 (34) | .560 | 1 | - |
| TWE | 93.00 (16.13) | -0.5 | -2.57 (34) | .015 | 5.8 | Moderate |

Game logs indicated generally good compliance with the assigned training schedule (Table 2). Variation in session durations resulted, as intended, in a normal distribution of TTT over the three groups (Shapiro-Wilk statistic=.966(35), *p*=.347, upper *BFB* =1).

**Table 2:** Compliance with assigned RECOGNeyes training protocol.

|  | Number of times/week instructed to play: |  |  |
| --- | --- | --- | --- |
|  | 2 times<br>(N=11) | 3 times<br>(N=12) | 4 times<br>(N=12) |
| Number of days actually played over two weeks | 4.3 (0.8) | 5.6 (1.9) | 6.2 (2.1) |
| Mean minutes played per day | 22.2 (3.8) | 23.9 (5.3) | 24.8 (8.5) |
| Mean gap in days between playing days | 3.6 (0.8) | 3.1 (1.4) | 2.7 (1.9) |
| Total minutes game played over two weeks | 94.1 (22.8) | 137.8 (64.1) | 152.5 (61.7) |

There was no evidence of reduced mental wellbeing after training. GHQ12 scores after training were lower (indicating better mental wellbeing) by an average of 1.3 (*SD*=3.8), *p*=.050, upper *BFB*=2.5.

There was no evidence of any adverse impact of assigned training schedule on change scores *F*<1, *p*=.827, upper *BFB* =1.

### Reading measures

Baseline values and changes in reading efficiency metrics are shown in Table 3. For all eye-movement metrics, a reduction is likely to indicate greater reading efficiency. Partial correlations between TTT and mean change on each metric, controlling for baseline performance, provided *weak* evidence that TTT was associated with improvement on three reading metrics, and *moderate* evidence for one metric, mean fixation duration. The upper BFB for the association between TTT and reduced fixation duration did not change substantially (upper *BFB*=4.9) when we also controlled for change in mean length in skipped words, suggesting that any impact of TTT on shorter fixation durations was not simply accounted for by hastier reading.

**Table 3:**
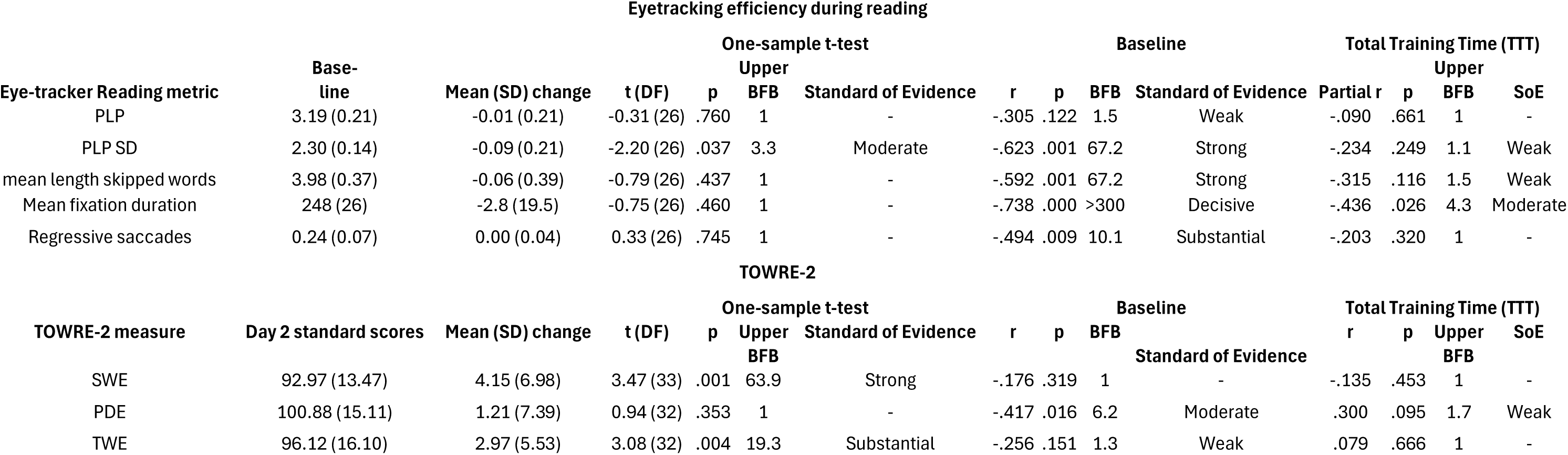
Changes in eye-tracking reading metrics, with effects of predictors (baseline values and TTT) on change.

### Antisaccade Task performance

Both speed and accuracy on the antisaccade task improved on Day 2 (Figure 2), more markedly for antisaccades than prosaccades. Overall accuracy as quantified by *d’* scores, was higher on Day 2 (*M*=3.57, SD=0.86) than on Day 1 (*M*= 4.09, *SD*=0.96), *F*(1,30)=11.251, *p*=.002, suggesting *strong* (upper *BFB*=29.6) evidence for a real increase. Antisaccade Cost was reduced on Day 2 (M=14.9, SD=22.7) compared to Day 1 (*M*=43.0, *SD*=32.9), *F*(1,30)=15,298, *p*=<.001, upper *BFB*=99.0, suggesting *very strong* (upper *BFB*=99.0) evidence for a real reduction in antisaccade cost.

**Figure 1:**
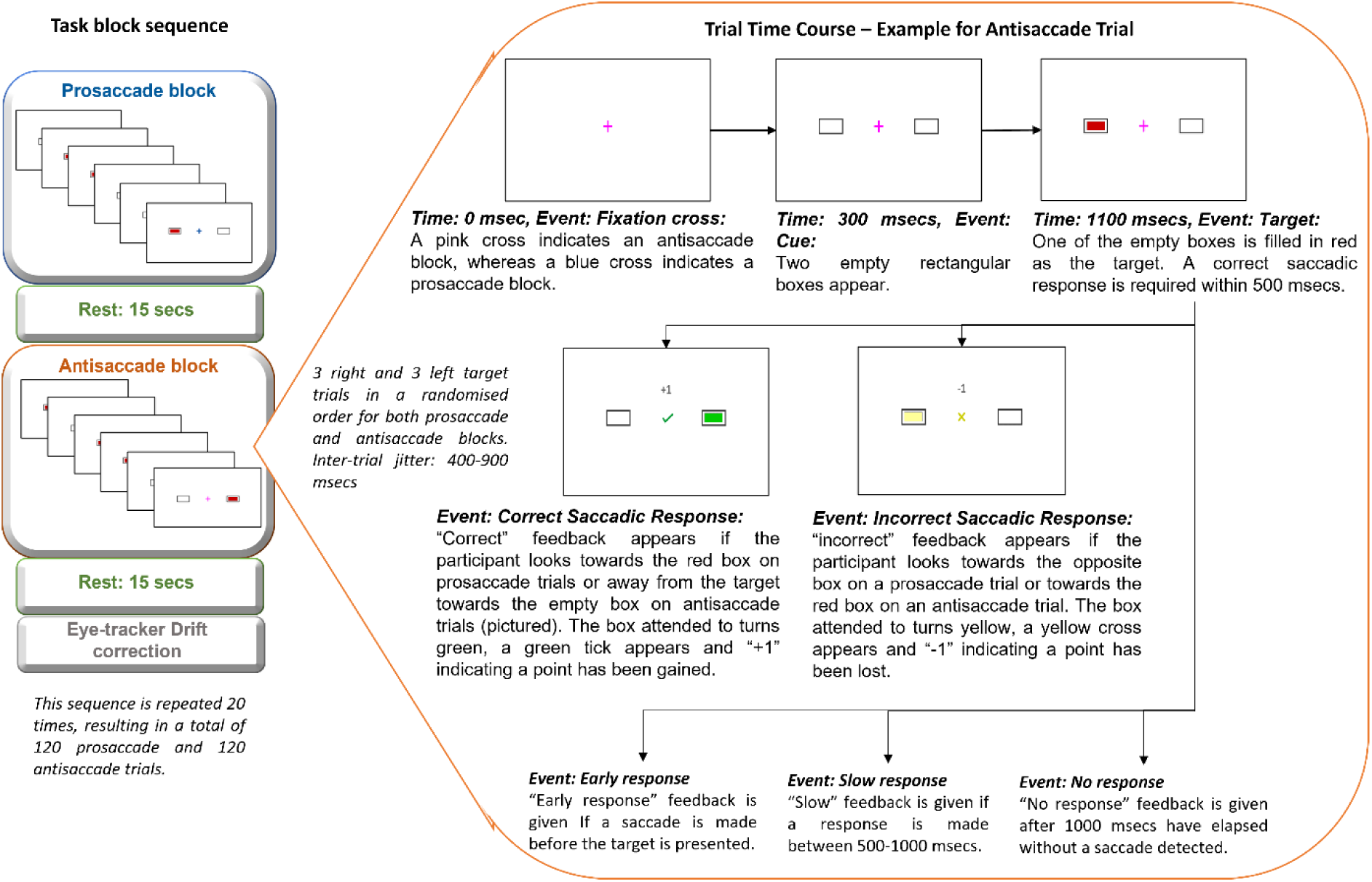
Pro/antisaccade task design.

**Figure 2:**
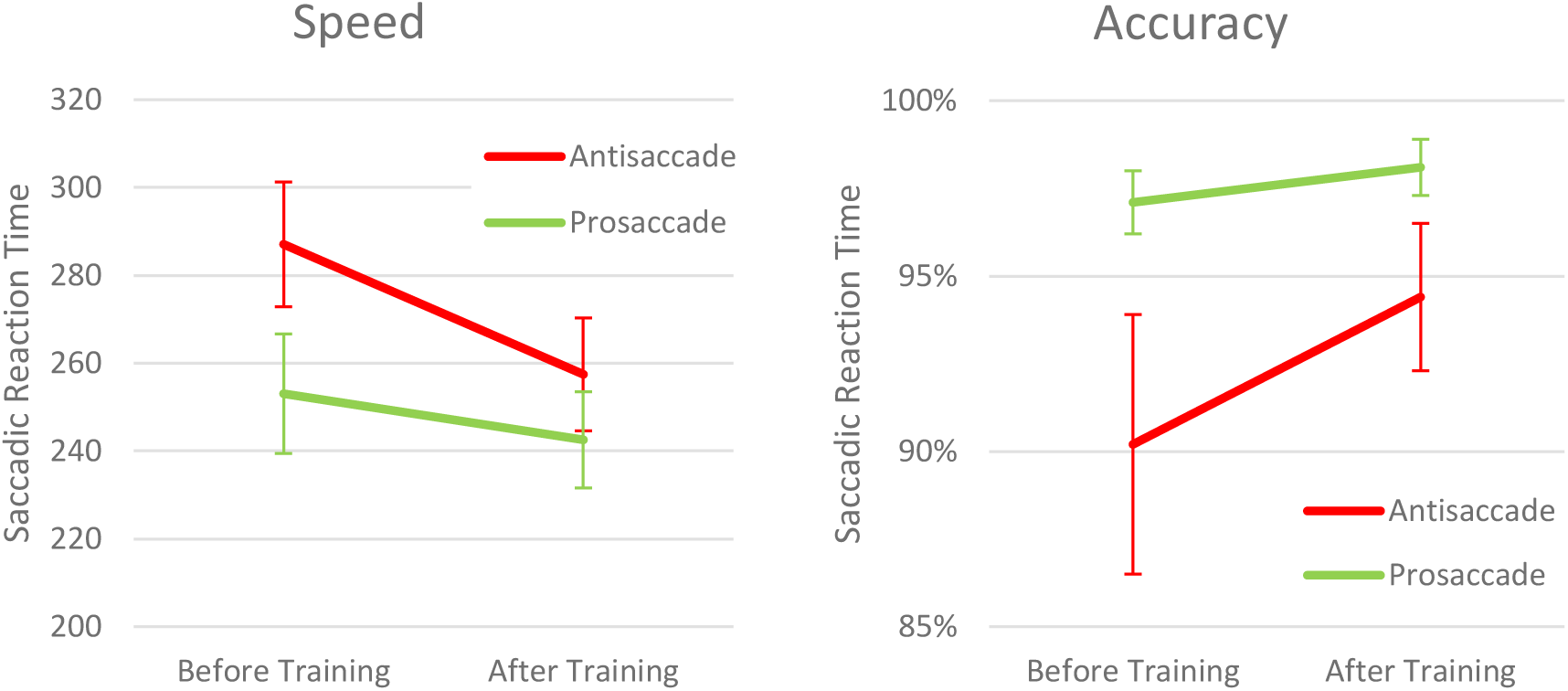
Changes in performance on each trial type before and after RECOGNeyes training.

Controlling for baseline performance, greater TTT was only *weakly* (upper *BFB*=1.3) associated with greater increases in *d’* scores, partial *r*= .27, *p*=.155, but *moderately* (upper BFB=4.7) with greater reductions in Antisaccade Cost, partial *r* =-.422, *p*=.020.

### Pupillometry

#### Phasic

Phasic pupil dilation rates were similar across days and conditions for the cue-target period. However, there was *decisive* evidence that pupil dilation rates between target onset and 100ms later were higher for antisaccades than prosaccades, *F*(1,29)=27.67, *p*<0.001, upper *BFB>300*), and *moderate* evidence that both were higher after than before training, *F*(1,29)=5.78, *p*=.023 upper *BFB*=4.7. Within participants, there was *decisive* trial-by-trial evidence that greater pupil dilation rates both periods (cue-target; target-100ms) were associated with shorter SRTs, *F*(1,29)=162.93, p<.002, upper BFB>300; *decisive* evidence that these correlations were stronger for the post target period *F*(1,29)=67.4, p<.001 upper *BFB*>30; and *moderate* evidence for stronger correlations for antisaccades than prosaccades, *F*(1,29)=6.86, p=.014,upper *BFB*=6.9. There was no evidence of any impact of training on phasic pupil dilation.

#### Tonic

There was *moderate* evidence that median pupil size at cue onset was smaller for antisaccade trial blocks than for pro-saccade blocks, *F*(1,29)=5.18, *p*=.030. upper *BFB* = 3.8, indicating higher tonic arousal ANS activity during antisaccade trials. There was no evidence of any impact of training on tonic pupil size.

### MEG measures of oscillatory brain activity: FEF alpha and DLPFC beta

#### Anticipatory period

We found no evidence for differences in either beta or alpha power between pro and antisaccades in either FEF or DLPFC. In both ROIs, alpha amplitudes tended to be higher than the mean for the whole trial period, and to rise during the anticipatory period, while beta amplitudes, as well as theta and delta, tend to show desynchronisation relative to the trial mean (Figure 3). We found no evidence of coupling between FEF alpha and DLPFC beta in either trial type. There was no evidence of overall change between Day 1 and Day 2 in either beta or alpha in either FEF or DLPFC.

**Figure 3:**
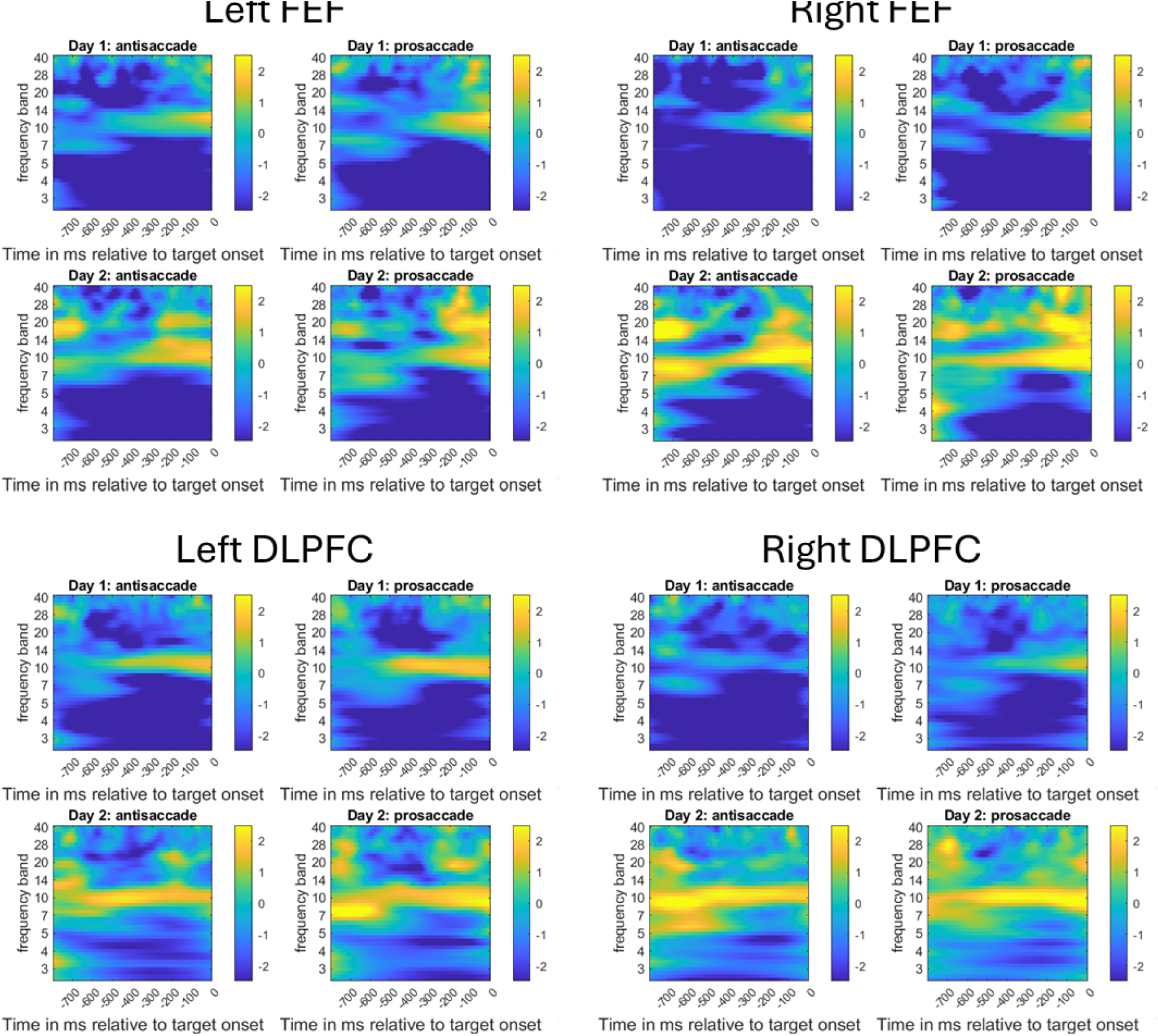
Oscillatory amplitudes for FEF and DLPFC ROIs during the anticipatory period by day and trial type. Colour values at each frequency band and time point in the panels represent t values for one-sample t-tests across participants, testing consistent deviation from each participant’s trial mean. Elevated alpha is apparent in all ROIs, tending to rise across the anticipatory period. There is also evidence of desynchronisation in both beta and lower frequency bands (theta and delta).

In left FEF, for antisaccade trials, we found *substantial* evidence (upper *BFB*=10.9) for an association between TTT and reduced amplitudes in a broadband cluster, centred on alpha but extending into beta and theta, in the latter part of the anticipatory period (Figure 4) trials. As this cluster maps on to frequencies that tend to rise towards the end of the anticipatory period (Figure 3, upper left), this implies that for antisaccade trials, greater TTT is associated with a greater reduction in the rate at which alpha and adjacent frequencies in left FEF rise as Target onset time approaches.

**Figure 4:**
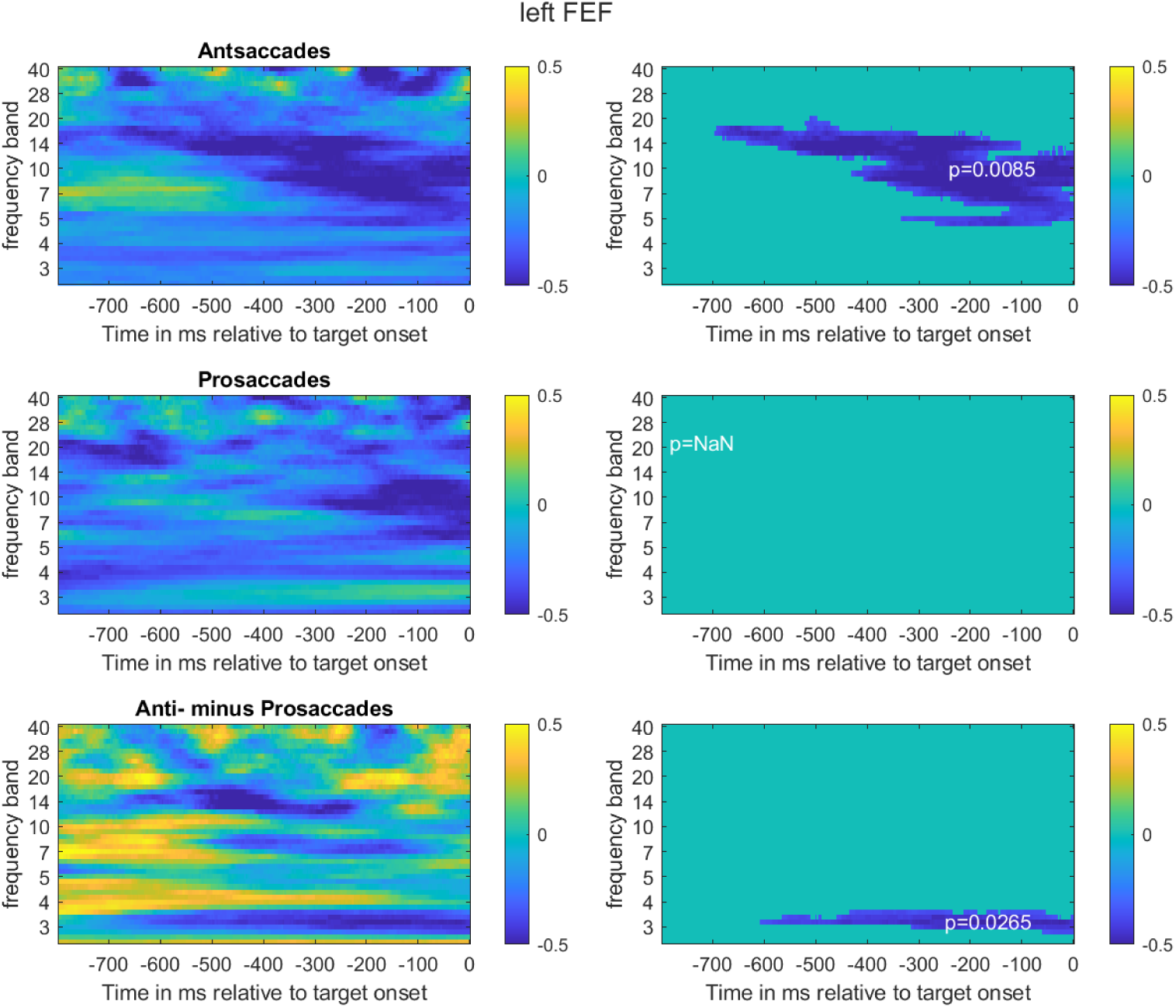
Partial correlations (rho values) between TTT and oscillatory amplitudes at each frequency band and time point for left FEF, controlling for baseline amplitudes. Right hand panels show results of tests for clusters of frequency/time points with rho-values satisfying p<.05. TTT was associated a cluster of amplitude reductions centred on alpha, but extending into the beta and theta range. TTT was also associated with greater increase (or smaller reduction) in delta amplitudes for prosaccade than antisaccade trial.

In both left and right DLPFC, greater TTT was associated with a greater beta desynchronisation for antisaccade trials (Figure 5). In left DLPFC, this evidence was *substantial* (upper *BFB*=26.5), and there was also *substantial* evidence (upper *BFB*<=12.8) for an association between greater TTT and a reduced rate of rise in alpha for prosaccades. In right DLPFC, for antisaccades, there was *decisive* (upper *BFB*>300) evidence that TTT was associated with reduced amplitudes in a cluster of beta and alpha frequencies, as well as with an unpredicted cluster of amplitude reductions in the theta range towards the end of the anticipatory period (upper *BFB*=4.9).

**Figure 5:**
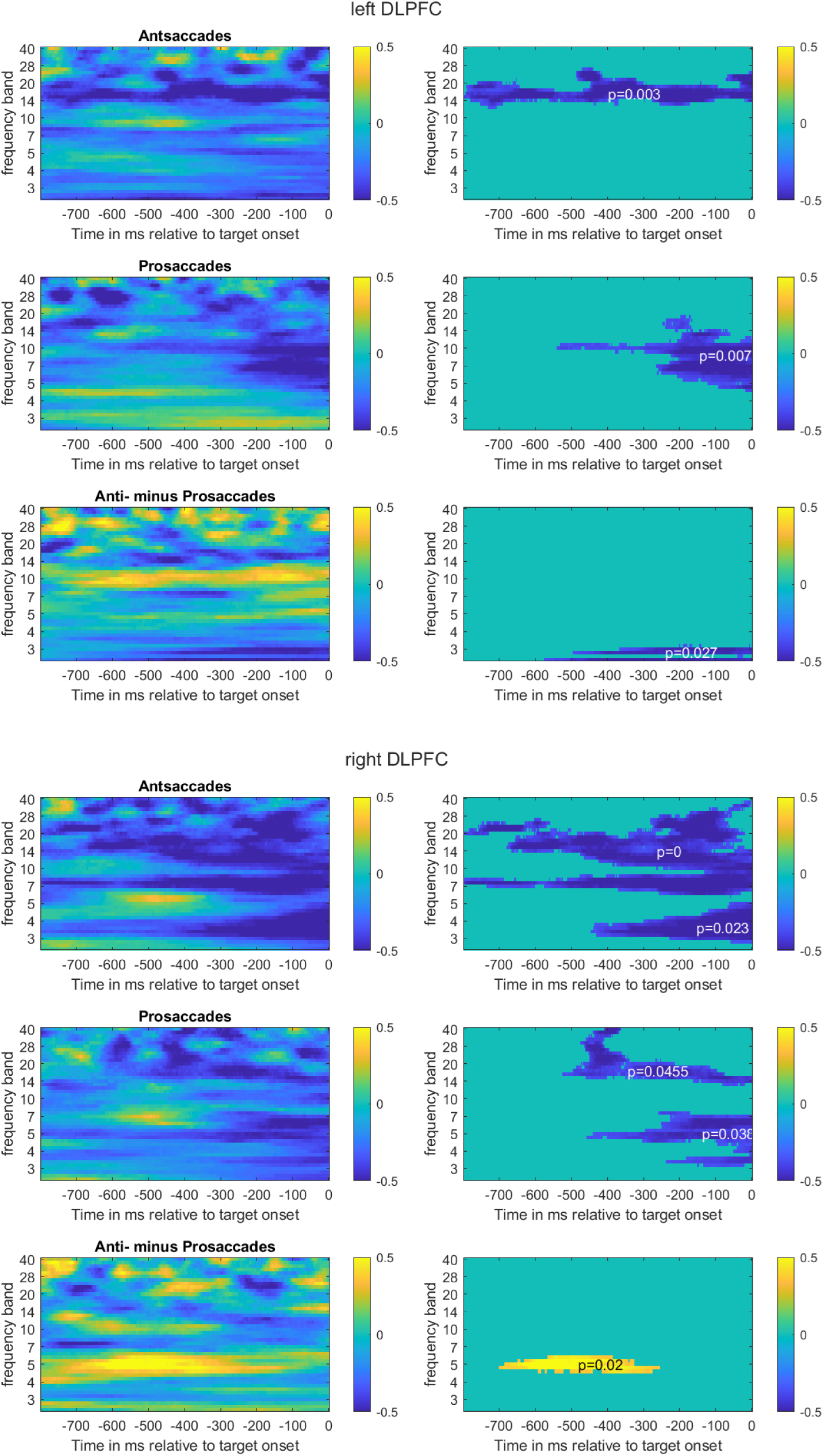
Partial correlations (rho values) between TTT and oscillatory amplitudes at each frequency band and time point for left and right DLPFC. Right hand panels show results of tests for clusters of frequency/time points with rho-values satisfying p<.05.

Similar patterns of correlations were found between anticipatory amplitude changes and improvements in Antisaccade Cost. To test whether the amplitude changes associated with greater TTT were correlated with amplitude changes associated with Antisaccade Cost, we conducted a resampling test in which, for each of 1000 iterations, we randomly selected 1% of the time frequency amplitude-change cells and computed the correlation between their Fisher-transformed *rho* values with TTT and with Antisaccade Cost respectively. Over the four ROIs (left and right FEF and DLPFC), the mean correlation was at least *r*=.66 for both antisaccades and prosaccades, thus supporting the interpretation that the variance in amplitude change associated with greater TTT was shared with variance in reduction in Antisaccade Cost.

#### Post target

Target-locked TFS plots indicated an increase in event-related PMBR from about 600 -1000ms post stimulus in ipsilateral FEF (Figure 6, left panel), with *moderate* (upper *BFB*=3.5) that for antisaccades, greater PMBR following RECOGNeyes training than at baseline. There was also *substantial* evidence (upper *BFB*<=23.1) that greater TTT was associated with greater PMBR increase (Figure 6, right panel). The timing (<600ms after Target onset) and ipsilateral location of this beta rebound suggests that it may represent a PMBR following the return saccade back to the central fixation point, rather than completion of the outward saccade. We found no evidence of PMBR time-locked to the outward saccade onset.

**Figure 6:**
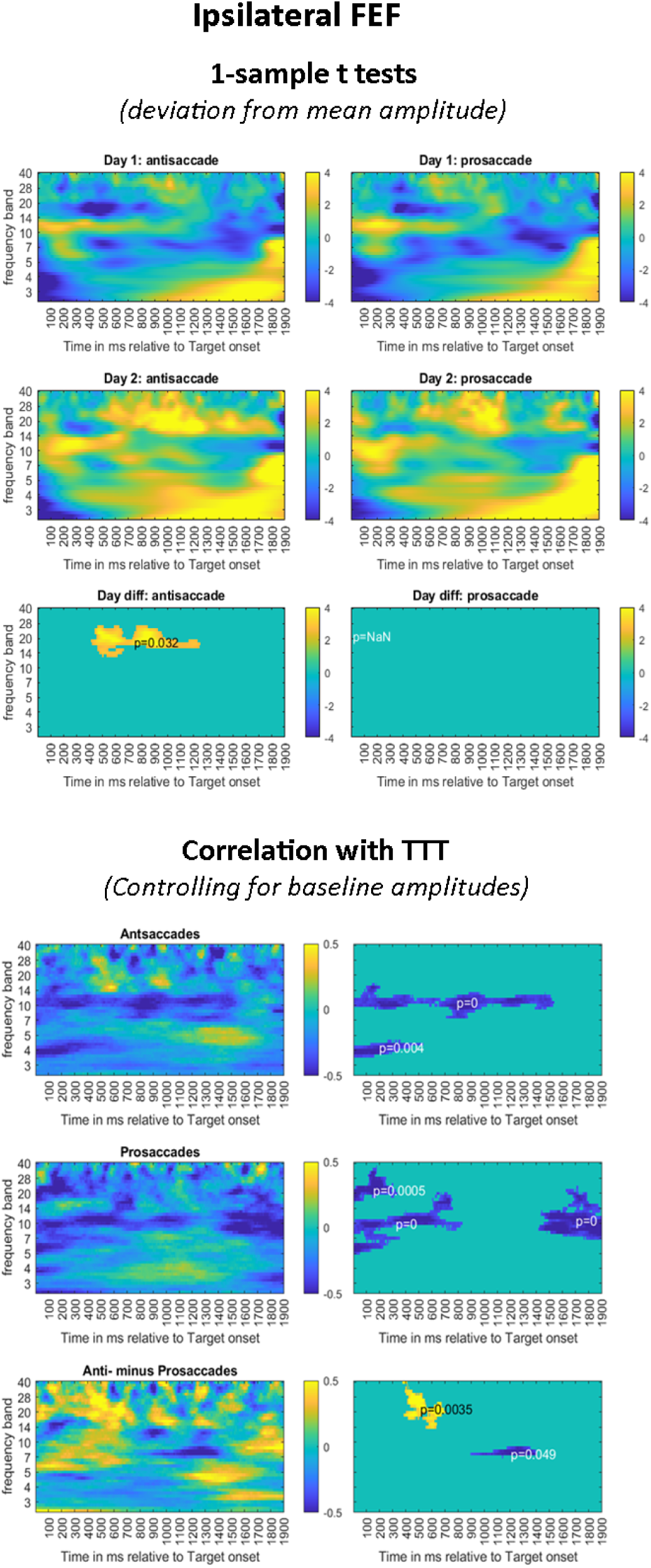
Upper panel : Evidence of PMBR beginning 600-1000ms post target in ipsilateral FEF. The cluster in the bottom right panel indicates PMBR on Day 2 for antisaccades. Lower panel: Partial correlations (controlling for baseline oscillatory amplitudes) between TTT oscillatory amplitudes at each frequency band and time point for ipsilateral FEF, indicating that greater training intensity was associated with greater PMBR.

### Resting state MR

#### Resting state fMRI connectivity analysis of the visual attention network

For homotopic ROI pairs, there was *moderate* evidence for a main effect of TTT *F*(1, 31) = 7.188, *p* = .012, upper *BFB*=7.8, indicating that greater TTT was associated with reduced homotopic connectivity across all homotopic ROI pairs. There was also *moderate* evidence of a TTT by hemisphere interaction, *F*(1, 31) = 5.509, *p* = .025, upper *BFB*=4.4 that indicated that greater TTT was associated with greater increase in intrahemispheric connectivity in left hemisphere relative to the right (Figure 7C). As there was *moderate* evidence that baseline intrahemispheric connectivity values were greater in the right hemisphere than in the left, *F*(1,33)=6.740, *p*=.014, upper *BFB*=6.9, the observed relationship between changes in intrahemispheric connectivity and TTT represents a change in the direction of more similar intrahemispheric connectivity in the two hemispheres.

**Figure 7:**
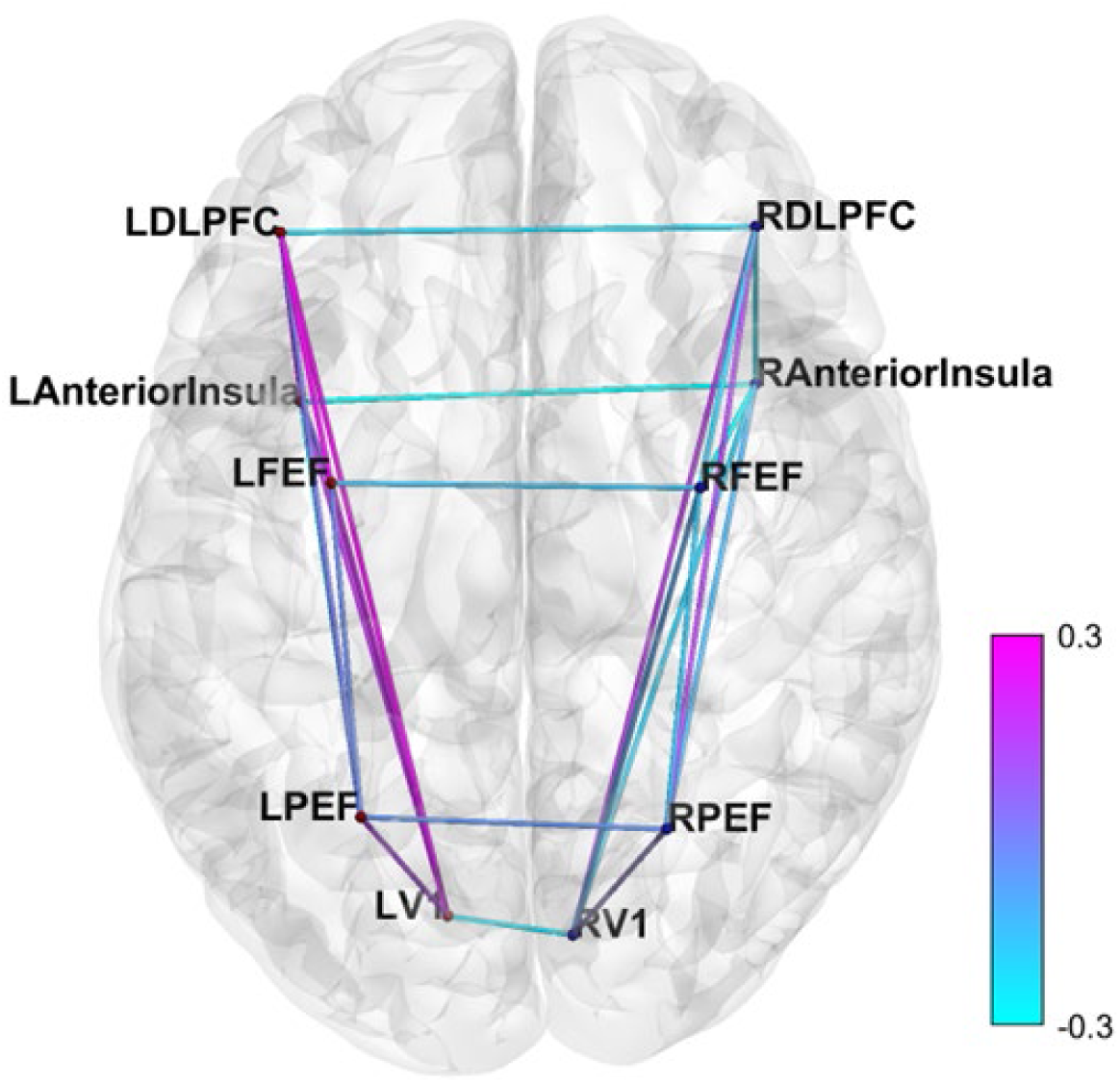
Resting state fMRI connectivity between oculomotor network ROIs. Greater TTT was associated with reduced homotopic connectivity and increased within-hemisphere connectivity in left hemisphere relative to right. Colour scale indicates strength of correlation between TTT and baseline-adjusted changes in connectivity. Bluer lines: Greater TTT associated with reduced connectivity; Pinker lines: Greater TTT associated with increased connectivity: ROIs (MNI coordinates): L DLPFC (-39.5, 37.5, 26.5); R DLPFC (40.5, 38.5, 25); L Anterior Insula (-36, 9, 4); Right Anterior Insula (40.5, 12, 0); L FEF (-31, -4.7, 50.5); R FEF (31.15, -5.5, 50.45); (L PEF (-26, -61, 55); R PEF (25.5, -63, 57.5); left primary visual cortex (-11.4, -77.7, 6,1)); right primary visual cortex (9.7, - 80.1, 5.8). V1=Primary visual Cortex. For MEG, the two V1 ROIs were replaced by a single ROI placed midway between LV1 and LV1 (-0.8, -79.3, 5.9).

## Discussion

After RECOGNeyes training, participants overall showed improvements in antisaccade task performance, as measured by greater accuracy and reduced Antisaccade Cost, greater Antisaccade Cost reductions being associated with more intensive RECOGNeyes training.

More intensive RECOGNeyes training was also associated with brain oscillatory changes in the pre-target anticipatory period, and these changes were also associated with reduced Antisaccade Cost. However, contrary to our prediction, the changes included decreases, rather than increases, in oscillatory alpha and beta power during the anticipatory cue-target interval, and in both antisaccade and prosaccade trials, and in both FEF and DLPFC, suggesting reduced anticipatory motor and attentional disinhibition (20,21,23,24) rather than inhibition. Nonetheless, given that more intensive training was associated with greater reductions in Antisaccade Cost without loss of directional accuracy, and that reductions in Antisaccade Cost were associated with similar changes in oscillatory amplitudes, this finding may indicate that, rather than improving performance by increasing inhibition prior to antisaccade trials, RECOGNeyes training reduces antisaccade cost by increasing attentional and motor readiness in both trial types, leading to increased antisaccade efficiency. This would be consistent with the finding that during reading, greater training intensity was associated with greater reduction in fixation times without an increase in the number of skipped words. The finding that greater training intensity was associated with greater antisaccade PMBR increase in FEF may reflect greater confidence in the predictive saccadic model, which would be consistent with greater antisaccadic efficiency.

The finding that at rest, greater training intensity was associated with reduced homotopic connectivity between oculomotor ROIS, and a relative increase in within-hemisphere connectivity in left hemisphere compared with the right was not an *a priori* prediction, but would be consistent with evidence from Jiang et al. (35) of reduced homotopic connectivity in children with ADHD, and may suggest that RECOGNeyes training may result in at least temporary plastic changes in the oculomotor network in the direction of greater independence of the left hemisphere network from the right. While a firm conclusion would require replication in a larger study, the finding provides some confidence that RECOGNeyes training may not simply impact neural processes transiently involved in execution of a trained oculomotor task, but also on more sustained functional connectivity patterns in the oculomotor network observable during rest.

We found no evidence of coupling between DLPFC beta and FEF alpha, as found by Hwang et al (20,54). However, our cue-target interval was considerably shorter (800ms) than that used in the Hwang task (1500ms) and may therefore index late-stage preparatory processing in both trial types.

We found no evidence that RECOGNeyes training impacted tonic pupil size or phasic pupil dilation rates. However, our finding that greater pupil dilation rate on a trial was associated with reduced SRT indicates that phasic arousal may be a useful additional training target to improve oculomotor efficiency.

## Conclusions and future directions

As this was an exploratory study any conclusions must be tentative, and additional exploration of the MEG and resting state data may be informative. However, the findings reported here offer some confidence in the concept of gaze-control training to improve inhibitory oculomotor control in inattentive young adults. Further research is required to establish whether training gains transfer to real-life improvements in cognitive control, produce worthwhile benefits in real-life tasks such as reading and written work, and whether gaze-control training games like RECOGNeyes could help younger children with ADHD and related neurodevelopmental conditions to acquire the oculomotor and cognitive control skills that underpin academic progress.

## Data Availability

All data produced in the present study are available upon reasonable request to the authors

