## Supplementary Materials for "Gaze-control training in a sample of inattentive young adults: A Confidence-in-Concept study of neural mechanisms"

**Pupillometry data cleaning methods**

Pupil size was recorded as the number of pixels within the detected pupil region area in arbitrary units (SR Research, 2017). Pupillometry data were cleaned using an in-house interactive MatLab script, with semi-automated detection and removal of blink artifacts. Extreme samples ±3 Std. Dev. from the trial mean were automatically excluded (1). Missing data periods were interpolated using spline interpolation (pchip). Data series were smoothed using a $4^{th}$-order low-pass Butterworth filter with 4Hz cut-off frequency. Trials were excluded if >30% samples were removed/missing (2,3).

Mean pupil diameter within a 200ms period 250 to 50 ms pre-cue was used as baseline, and pupil sizes expressed as a proportion of this mean diameter (4). Rate of pupil dilation was computed (5) by taking the difference in pupil size at each event and dividing this by the duration of this period of interest.
